# Development of Prognostic Models for Survival and Care Status in Sporadic Creutzfeldt-Jakob disease

**DOI:** 10.1101/2022.02.06.22270293

**Authors:** Akın Nihat, Janice M. Ranson, Dominique Harris, Kirsty McNiven, Peter Rudge, John Collinge, David J. Llewellyn, Simon Mead

**Affiliations:** MRC Prion Unit at UCL, UCL Institute of Prion Diseases, Cleveland St, London; National Prion Clinic, National Hospital for Neurology and Neurosurgery, University College London Hospitals NHS Foundation Trust, London; College of Medicine and Health, University of Exeter, Exeter; Deep Dementia Phenotyping Network; Alan Turing Institute, London

## Abstract

Sporadic Creutzfeldt-Jakob disease (sCJD), the most common human prion disease, typically presents as a rapidly progressive dementia and has a highly variable prognosis. Despite this heterogeneity, clinicians need to give timely advice on likely prognosis and care needs. No prognostic models have been developed that predict survival or time to increased care status from the point of diagnosis. We aimed to develop clinically useful prognostic models with data from a large prospective observational cohort study. 537 patients were visited by mobile teams of doctors and nurses from the NHS National Prion Clinic within 5 days of notification of a suspected diagnosis of sCJD, enrolled to the study between October 2008 and March 2020, and followed up until November 2020. Prediction of survival over 10-, 30- and 100-day periods was the main outcome. Escalation of care status over the same time periods was a secondary outcome for a subsample of 113 patients with low care status at initial assessment. 280 (52.1%) patients were female and the median age was 67.2 (IQR 10.5) years. Median survival from initial assessment was 24 days (range 0-1633); 414 patients died within 100 days (77%). Ten variables were included in the final prediction models: sex; days since symptom onset; baseline care status; *PRNP* codon 129 genotype; MRC (Medical Research Council) Prion Disease Rating Scale, Motor and Cognitive Examination Scales; count of MRI abnormalities; Mini-Mental State Examination score and categorical sCJD phenotype. The strongest predictor was *PRNP* codon 129 genotype (odds ratio 6.65 for MM compared to MV polymorphism; 95% CI 3.02-14.68 for 30-day mortality). Of 113 patients with lower care status at initial assessment, 88 (78%) had escalated care status within 100 days, with a median of 35 days. Area under the curve for models predicting outcomes within 10, 30 and 100 days was 0.94, 0.92 and 0.91 for survival, and 0.87, 0.87 and 0.95 for care status escalation respectively. Models without *PRNP* codon 129 genotype, which is not immediately available at initial assessment, were also highly accurate. We have developed a model that can accurately predict survival and care status escalation in sCJD patients using clinical, imaging and genetic data routinely available in a tertiary national referral service. The utility and generalizability of these models to other settings could be prospectively evaluated when recruiting to clinical trials and providing clinical care.

## Introduction

Sporadic Creutzfeldt-Jakob Disease (sCJD) is a fatal neurodegenerative disease that manifests as a usually rapid, multidomain dementia associated with myoclonus, motor, and sensory impairments. Median survival from symptom onset is five months^1^. Prognosis is highly variable and right-skewed in distribution, ranging from a few weeks to over ten years^2,3^. Whilst there is no treatment to cure or slow disease progression, appropriate patient management involves the avoidance of needless investigations and timely care planning^4,5^. In the UK, patients are often investigated and diagnosed during a hospital admission. Potential discharge options include hospice care, another 24-hour care facility, home care-package, or no/informal care at home. These decisions are informed by clinical judgement of likely prognosis and the speed of clinical decline anticipated. Accurate prognostication and prediction of care status are therefore crucial for provision of optimal and timely support.

Human prion diseases, of which sCJD is the most common, involve the templated misfolding of cellular prion protein into disease-associated assemblies, including forms that propagate by fission and forms that cause neurodegeneration^6^. Prion diseases are associated with propagation of prion strains, which lead to distinct clinical and pathological phenotypes which are maintained on transmission to other humans or animals and are encoded by the molecular structure of prions^6^. The prion protein gene (*PRNP*) is polymorphic in many populations, with a common variation being found at codon 129, encoding amino acids methionine or valine. Numerous clinicopathological and molecular studies have shown the codon 129 genotype as a key determinant of survival, rate of progression and clinical phenotype, in part related to conformational selection of permissible prion strains^1,7,8^. Several demographic, protein biofluid, brain imaging, and neurophysiological biomarkers are associated with prognosis or subtypes of sCJD^1,9–14^. We and others have evaluated several tools to measure patient progression^3,15^. This knowledge provides a rational basis to improve prognostication using multivariable modelling.

Previous studies have had some success in predicting survival but have significant limitations. Staffaroni *et al*. identified cerebrospinal fluid (CSF) biomarkers associated with survival^16^ and Sundaram *et al*. used a neuropsychological assessment score to predict probability of three-, six- and twelve-month survival^17^ though neither tested combinations of predictors. Llorens *et al*. developed a prognostic model of 6-month survival from sCJD symptom onset incorporating age, sex, *PRNP* codon 129 genotype, and CSF tau, which whilst valuable, did not model survival from the point of clinical decision making. Model performance was moderate (area under the curve=0.686; 95% confidence interval, 0.67–0.71), and they did not produce a prognostic model for escalating care status^18^. Here, we aimed to develop accurate individual prognostic models for survival from the point of a clinical assessment, and separately model care need progression using a wide range of prospective clinical, imaging, genetic and biomarker data from the UK’s National Prion Monitoring Cohort study.

## Methods

Reporting of our study follows the Transparent Reporting of a multivariable prediction model for Individual Prognosis or Diagnosis (TRIPOD) statement^19^ (see the TRIPOD checklist in eMethods).

### Data and study population

We used longitudinal data on 537 patients with sCJD from the National Prion Monitoring Cohort (Cohort study) described in detail elsewhere^3^. The sample comprised all patients with sCJD enrolled in the Cohort study in March 2020. The Cohort study is a UK-wide observational study of human prion diseases led by the NHS National Prion Clinic, which prospectively collects data from patients as part of a clinical assessment including demographics, key investigation results, imaging, prion protein gene sequencing and bespoke clinimetric scales^3,15^. The Cohort study received ethical approval from Scotland MREC A.

A national referral system for prion diseases was set up in the UK in 2004. UK neurologists were asked by the Chief Medical Officer to refer all patients with suspected prion disease jointly to the National CJD Research and Surveillance Unit (Edinburgh, UK) and to the NHS National Prion Clinic (London, UK) for the purpose of both epidemiological surveillance, provision of specialist clinical care and also participation in clinical research, including clinical trials and the Cohort study. The Cohort study began in October 2008, and aimed to enrol all symptomatic patients with prion disease in the UK thereafter. The Cohort study collected natural history data very similar to that used in the MRC-PRION1 trial of quinacrine 2004-2008, and merged with the natural history data from this trial^20^. In 2015, in order to focus on the natural history of patients who were not already moribund, the protocol was altered such that patients with high levels of neurodisability and MRC Scale <5 at initial assessment were no longer eligible. Not all cases of sCJD are diagnosed and referred in life, but over the study period this was certainly >90%. The Cohort study team visit patients within 5 working days of notification. For the purposes of this report, the initial study visit was assumed to be the point of diagnosis. In reality, it is difficult to define a precise point of diagnosis of sCJD as evidence typically accrues over a period of a few weeks in-patient care, based on a combination of the clinical features and results of MRI brain, CSF and EEG investigations. Patients were diagnosed according to contemporary diagnostic criteria with very high specificity^21^. In the absence of disease-modifying therapeutics, only supportive symptomatic treatment, for example treatments for psychiatric disturbance, myoclonus, sleep disorder, and end of life care, was given. Post-mortem examination was conducted in 241 of 504 (48%) patients who died during follow-up. Diagnostic accuracy was confirmed in all cases.

### Outcome variables

#### Survival

The primary outcome variable was death (yes / no) by three time-points: 10-days, 30-days and 100-days from first assessment. These outcomes were decided based on an expectation of judgements to be made by the treating physician: 10 day survival implies immediate end-of-life care, in the UK typically hospice care; whereas 100 day survival implies informal care, set up of an external care package at home, or consideration of nursing home care.

#### Increased care status

The secondary outcome variable was increased care status (yes / no), investigated in a subsample of 113 patients who at the time of initial assessment had lower care status (either no care, informal care (for example, some support from a family member), or formal care up to two times daily) and had a date recorded for progression to increased care status (formal care more than twice daily, or nursing home / hospice placement). This secondary outcome was assessed at the same time-points as the primary outcome: 10-days, 30-days and 100-days from first assessment.

### Predictor variables

A scoping literature review and expert panel discussion identified 29 variables associated or potentially associated with prion disease progression or prognosis. The review prioritised research evidence of an association between a predictor and CJD survival, rather than factors known to influence dementia survival more generally^22^, and we excluded variables that are not feasibly available at diagnosis (serum tau, serum neurofilament light, prion molecular strain type [combination of *PRNP* codon 129 genotype and western blot type]), and those symptoms that may be a very significant burden for carers but fluctuate over short periods of time, or are treatable (eg. myoclonus, psychiatric disturbance); 25 potential predictors remained. To reduce this further to minimise multiple testing, the list of variables was independently rated by two clinicians with sCJD assessment expertise (AN and SM, see eMethods 2), resulting in 13 candidate predictors for analysis.

#### Sociodemographics

Sex (male/female), age (years), and days since symptom onset were based on patient or relative reports.

#### Clinical assessments

The MRC Prion Disease Rating Scale score (MRC Scale; 0-20)^3^ is a functional composite measure of sCJD disease progression developed using item-response modelling. The Prion Disease Motor Scale (0-100)^15^ and Cognitive Scale (0-100)^15^ were later developed to measure progression of motor and cognitive impairments using a similar methodology. Mini-Mental State Examination (MMSE) total score (0-30)^23^ was also recorded. The clinical phenotype of sCJD was based on clinical judgement of the dominant symptomatic presentation as one of 7 categories; ‘visual’, ‘ataxic’, ‘cognitive’, ‘psychiatric/behavioural’, ‘sleep/thalamic’, ‘stroke-like’ and ‘classical’^24^. Due to the small number of patients in some categories we collapsed this to a 3-category clinical phenotype variable of ‘classical’, ‘cognitive, psychiatric or behavioural’, and ‘other’. Care status was recorded for all assessments based on the following scale: admission to nursing home or hospice (0), formal care three/four times per day (“all care”) (1), formal care up to twice per day (2), informal care (e.g. spouse, other relative or friend, not local authority or hospital care) (3), or no care (4). Data were collected between October 2008 and November 2020, with a total follow-up of 161 patient-years. For the primary analyses, due to the small number of patients in some of the care status categories we combined categories 1, 2, 3 and 4 to produce a binary variable of ‘nursing home or hospice care’ (yes/no). The secondary analyses predicting care status progression to category 0 or 1 (higher care status), was conducted using data on 113 patients with baseline care status category 2, 3 or 4 (lower care status). We used a binary variable representing formal care status at baseline (yes/ no), defined as category 2 versus category 3 or 4.

#### Imaging

Local diagnostic diffusion-weighted magnetic resonance imaging (MRI) already available at the time of first assessment was used to assess the presence of five abnormalities by the recruiting neurologist (each yes/no); atrophy, cortical ribboning, pulvinar sign, basal ganglia and thalamic). These were modelled as a single count variable (possible range 0 – 5). Necessarily this involved a range of scanner manufacturers and protocols as only a small proportion of sCJD patients are able to transfer to a regional or national specialist site for research investigations; given the wide geographical distribution of sCJD patients throughout the UK and absence of phenotypic clusters, we reasoned that any variation in imaging quality or protocol between referring hospitals would be random, and not bias results.

#### Genetics

*PRNP* codon 129 genotype (MM, MV or VV) was determined by *PRNP* sequencing^25^. This variable was included as a candidate predictor as the genetic data is collected during the baseline assessment, via peripheral blood sample. However, it is worth noting that the results confirming *PRNP* codon 129 genotype currently take from several days to weeks to obtain in clinical practice.

#### Biomarkers

Presence of electroencephalography Periodic Sharp Wave Complexes (PSWCs) was determined by reference to the local neurophysiology report of electroencephalogram(s) (EEG) available at first assessment. Cerebrospinal fluid (CSF) s100b values were determined by local reports.

### Data analysis

All analyses were performed using STATA version 16.0, including installation of the additional packages: st0177, mim, mfpmi, st0569, diagt, missings, st0139_1, looclass.

#### Missing data

Multivariate imputation by chained equations was conducted for missing data on all predictors according to guidance^26^. Eighty imputed datasets were created, this number being greater than the percentage of cases with incomplete data (79%). Analyses were restricted to those with complete data on the outcome variables. Predictive mean matching to three nearest neighbours was used for continuous variables, multinomial logistic regression for categorical variables, ordinal logistic regression for ordered categorical variables, and logistic regression for binary variables. Variables were imputed iteratively, from the one with the least missing data to the most. Age at enrolment, sex and survival outcomes were included in the imputation model as complete auxiliary variables.

#### Primary outcome model development

For the primary outcome of survival, prediction models were developed using a multivariable fractional polynomial approach^27^. For each time-window, logistic regression using backwards elimination (BE) with a rejection criterion of p ≥ 0.05 was conducted on 1000 random bootstrap replication samples with replacement. In each replication, BE was combined with a closed test function selection procedure (FSP). FSP is a systematic search for non-linearity that investigates the most appropriate functional form for each continuous predictor, considering first- and second order fractional polynomial terms to four degrees of freedom. This procedure ensures that variables are not erroneously disregarded due to inappropriate modelling of non-linear associations. To avoid overfitting, linear terms are used to model associations between continuous predictors and the outcome unless a more complex term provides a significantly better fit (α = 0.05).

#### Selection of predictors

Predictors were selected based on the proportion of times they were selected by the BE regression model in the bootstrapped samples. To aid clinical utility, we selected a single set of predictors for use in the final models predicting 10-day, 30-day and 100-day mortality. Variables with a bootstrap inclusion fraction (BIF) of 50% or more resulting from the *mfpboot* analysis for any of the three time-points were selected for inclusion in all three final models. Sensitivity analyses were conducted comparing model fit with and without each of the borderline variables not selected for the final models (borderline variables were defined as having a BIF between 25% and 49%).

#### Secondary Outcome Model Development

Variables selected in the final model for the primary analyses were used to predict increased care status. Final models predicting increased care status were developed using the same approach as above, for the same three time-points: 10-days, 30-days and 100-days from first assessment.

#### Estimation of the final predictive models

Six multivariable fractional polynomial logistic regression models were estimated in the multiply imputed data, predicting 10-day, 30-day and 100-day mortality, and 10-day, 30-day and 100-day increased care status. The most appropriate functional form of continuous predictors was identified independently in each final model using the FSP described above. Predicted probabilities were calculated based on the model coefficients and averaged across imputations.

#### Evaluation of model performance

Receiver operating characteristic (ROC) curves and areas under the curve (AUC) were used to evaluate the discrimination performance of each of the 6 models. For each model, a binary classifier was generated using a predicted probability threshold of 0.5 (assuming equal importance of false positives and false negatives). This classifier was used to calculate rates of true-positives, true-negatives, false-positives and false-negatives, overall percentage correctly classified, and model performance statistics including sensitivity, specificity, positive predictive values (PPV) and negative predictive values (NPV). In absence of a suitable external validation dataset to estimate the generalisability of the models due to the rarity of sCJD we repeated the model evaluation using leave-one-out cross-validation. This procedure involves estimating the model on n-1 observations, and applying the resulting prediction to the excluded observation. This was repeated excluding each observation in the data, and results were combined to generate the overall cross-validated model performance statistics.

## Results

### Sample characteristics

Detailed characteristics of the clinical sample is shown in supplementary eTable 1. The total sample (N=537) had a median age of 67.16 (IQR 10.53), 280 (52.1%) were females and 198 (36.9%) were resident in a nursing home or hospice at baseline. The median delay between carer-reported symptom onset and date of the initial diagnostic assessment was 130.8 days. Median survival from initial assessment was 24 days (IQR = 71, range 0-1633). The number of patients who died within 10, 30 and 100 days from initial assessment was 127 (23.7%), 293 (54.6%) and 414 (77.1%) respectively. Of those with lower care status at baseline who progressed to requiring increased care (N = 113), the median time until increased care status was 35 days (IQR = 73, range 2-251). The number of patients requiring increased care status within 10, 30 and 100 days was 20 (17.7%), 55 (48.7%), and 88 (77.9%) respectively.

**Table 1.**
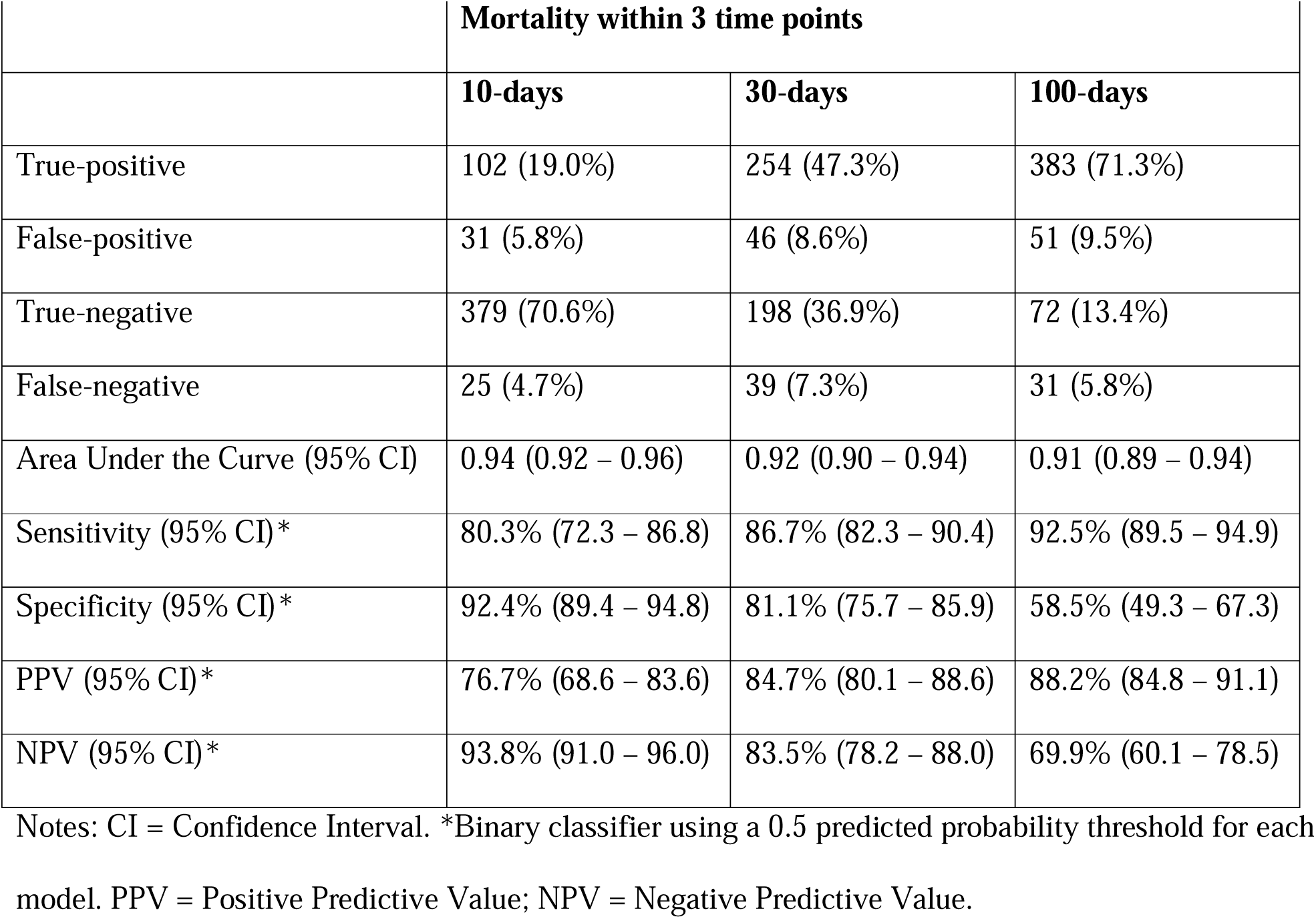
Classification rates and performance of models predicting 10- 30- and 100-day mortality in sCJD patients (N=537).

### Selection of predictors

Of the 13 candidate predictors, age, EEG and CSF s100b abnormality did not have a BIF≥ 50% in any of the three analyses and were not selected for inclusion in the final model. Final models comprised the ten remaining variables: sex; days since symptom onset; baseline care status; *PRNP* codon 129 genotype; MRC Prion Disease Rating, Motor and Cognitive Scales, count of MRI abnormalities; MMSE score and clinical phenotype. Of variables included in the final models, those with the highest proportion of missing data were clinical phenotype category (32.7% of patients), Cognitive score (26%) and codon 129 (18%); all other variables were missing for less than 7% of patients.

### Primary models predicting survival (N=537)

Final models predicting 10- 30- and 100-day mortality are detailed in Supplementary eTable 2. The overall percentage correctly classified by each model was 89.6%, 84.2% and 84.7% respectively. Classification rates and model performance statistics are presented in Table 1. All leave-one-out cross-validation results were within the 95% CI range estimated using the original development sample (see Supplementary eTable 3). ROC curves for the primary models predicting survival are presented in Figure 1a. The actual survival of patients stratified by model predictions and 10, 30 and 100 days is shown in Figure 2.

**Figure 1a.**
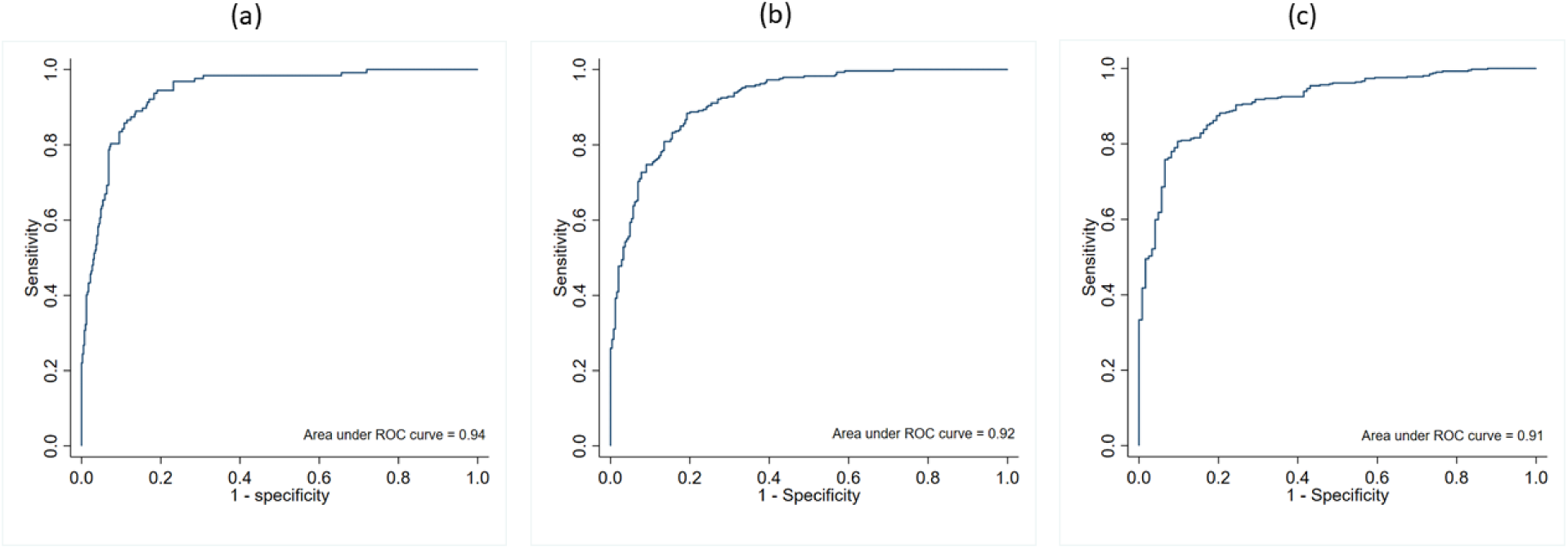
ROC curves for primary models (N=537) predicting mortality within (a) 10 days, (b) 30 days, and (c) 100 days.

**Figure 1b.**
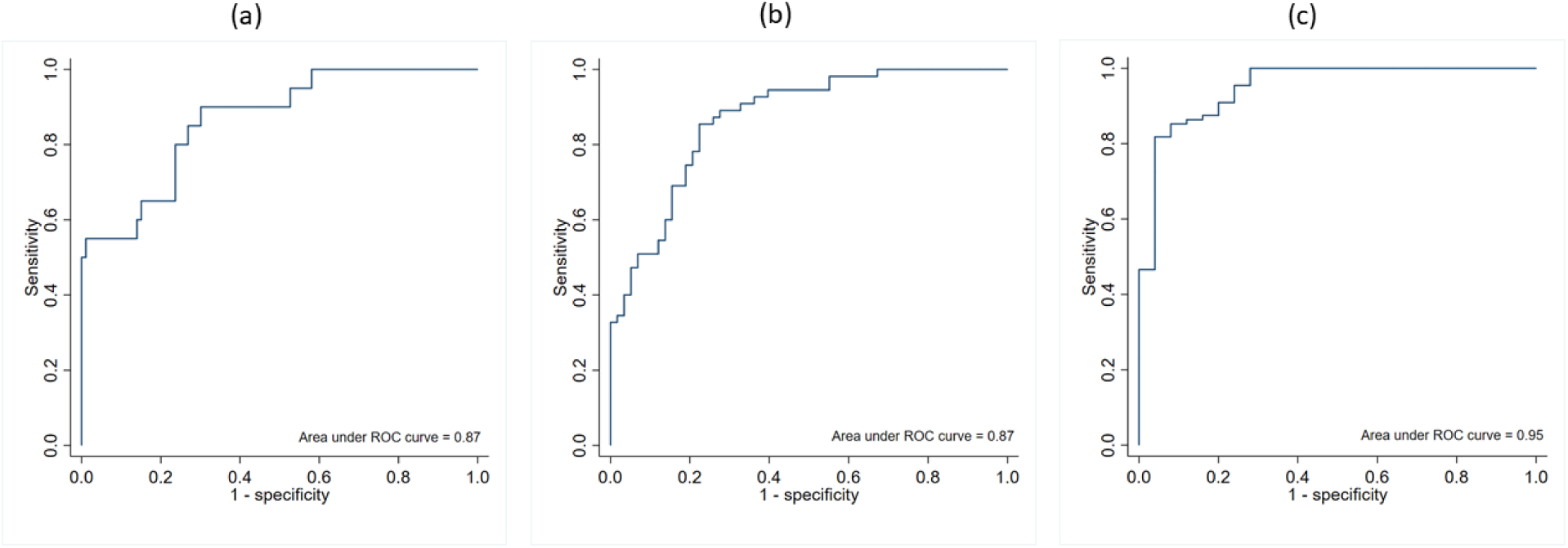
ROC curves for secondary models (N=113) predicting increased care status within (a) 10 days, (b) 30 days, and (c) 100 days.

**Table 2.** Classification rates and performance of models predicting 10- 30- and 100-day increased care status in sCJD patients with lower care status at baseline (N=113).

|  | **Progression to increased care status within 3 time points** |  |  |
| --- | --- | --- | --- |
|  | **10-days** | **30-days** | **100-days** |
| True-positive | 10 (8.9%) | 42 (37.2%) | 84 (74.3%) |
| False-positive | 1 (0.8%) | 12 (10.6%) | 7 (6.2%) |
| True-negative | 92 (81.4%) | 46 (40.7%) | 18 (15.9%) |
| False-negative | 10 (8.9%) | 13 (11.5%) | 4 (3.5%) |
| Area Under the Curve (95% CI) | 0.87 (0.78 – 0.95) | 0.87 (0.81 – 0.93) | 0.95 (0.90 – 1.00) |
| Sensitivity (95% CI)* | 50.0% (27.2 – 72.8) | 76.4% (63.0 – 86.8) | 95.5% (88.8 – 98.7) |
| Specificity (95% CI)* | 98.9% (94.2–100.0) | 79.3% (66.6 – 88.8) | 72.0% (50.6 – 87.9) |
| PPV (95% CI)* | 90.9% (58.7 – 99.8) | 77.8% (64.4 – 88.0) | 92.3% (84.8 – 96.9) |
| NPV (95% CI)* | 90.2% (82.7 – 95.2) | 78.0% (65.3 – 87.7) | 81.8% (59.7 – 94.8) |

Notes: CI = Confidence Interval. \*Binary classifier using a 0.5 predicted probability threshold for each model. PPV = Positive Predictive Value; NPV = Negative Predictive Value.

**Table 3.** Sensitivity analyses comparing the area under the curve of primary models and alternative models in predicting 10-day, 30-day and 100-day mortality in sCJD patients.

|  | **10-day mortality**<br>**AUC (95% CI)** | **30-day mortality**<br>**AUC (95% CI)** | **100-day**<br>**mortality**<br>**AUC (95% CI)** |
| --- | --- | --- | --- |
| Primary model | 0.94 (0.92 – 0.96) | 0.92 (0.90 – 0.94) | 0.91 (0.89 – 0.94) |
| Primary model plus EEG abnormality | 0.94 (0.92 – 0.96),<br>$p=0.152$ | 0.92 (0.90 – 0.94),<br>$p=0.282$ | 0.91 (0.89 – 0.94),<br>$p=0.061$ |
| Primary model plus CSF s100b abnormality | 0.94 (0.92 – 0.96),<br>$p=0.450$ | 0.92 (0.90 – 0.94),<br>$p=0.313$ | 0.91 (0.89 – 0.94),<br>$p=0.675$ |
| Primary model plus EEG and CSF s100b abnormality | 0.94 (0.92 – 0.97),<br>$p=0.073$ | 0.92 (0.90 – 0.94),<br>$p=0.498$ | 0.91 (0.89 – 0.94),<br>$p=0.439$ |
| Primary model without *PRNP* codon 129 genotype | 0.94 (0.91 – 0.96),<br>$p=0.190$ | 0.90 (0.88 – 0.93),<br>$p<0.001$ | 0.89 (0.86 – 0.92),<br>$p=0.001$ |

AUC = Area under the curve.

**Figure 2.**
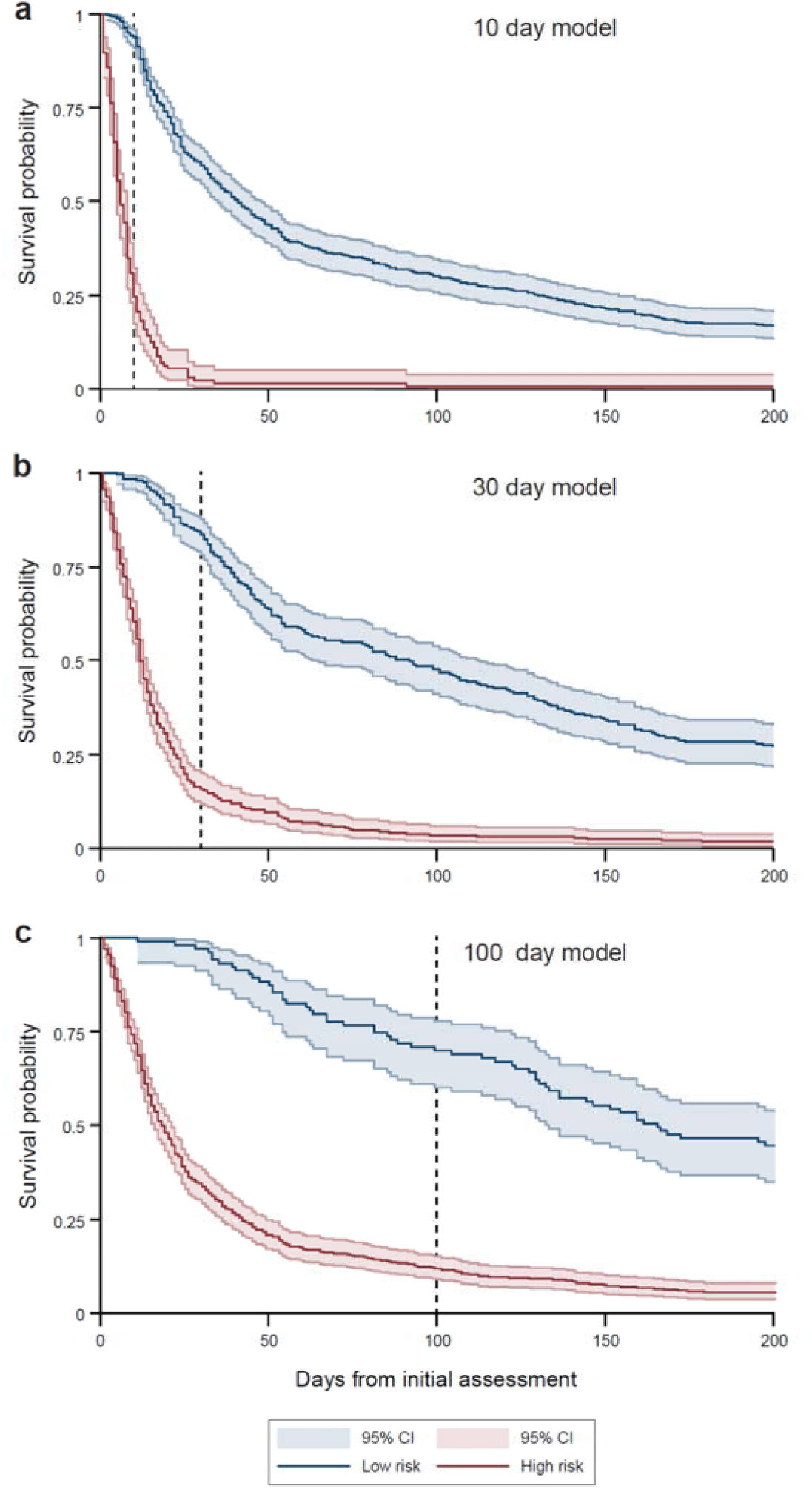
Actual survival of patients stratified by model prediction of death within 10, 30 and 100 days.

### Secondary models predicting increased care status (N=113)

Secondary models predicting 10- 30- and 100-day increased care status are detailed in Supplementary eTable 4. The overall percentage correctly classified by each model was 90.3%, 77.9% and 90.2% respectively. Classification rates and model performance statistics are presented in Table 2. All leave-one-out cross-validation results were within the 95% CI range estimated using the original development sample (see Supplementary eTable 5). ROC curves for secondary models predicting increased care status are presented in Figure 1b.

### Sensitivity analyses

Four sensitivity analyses were conducted in each of the three primary models predicting 10-, 30- and 100-day mortality. Two variables not initially selected for inclusion had borderline BIF (25%-49%). These were EEG and CSF s100b, with BIF of 43.5% and 40.6% respectively. We therefore estimated the primary models including these variables separately, and together, and used AUC to compare overall model performance with the primary model. As a fourth sensitivity analysis, we compared the overall model performance with and without *PRNP* codon 129 genotype. This variable was initially included in the final models, although results confirming *PRNP* codon 129 genotype are not currently available immediately, and so assessing the contribution of this variable to model performance could inform the importance of expediting these results. As shown in Table 3, none of the primary models were improved by the addition of EEG abnormality, CSF s100b abnormality, or both variables together. Exclusion of *PRNP* codon 129 genotype significantly reduced performance of models predicting 30-day and 100-day mortality (although the models still performed well, AUC=0.90 and 0.89 respectively).

## Discussion

We have developed accurate prognostic models for survival and care status escalation in sCJD patients using data routinely available in tertiary diagnostic services. We observed highly heterogenous outcomes for both survival and care status, reinforcing the need for accurate prognostic decision-making aids to enhance clinical trials and clinical care. Whilst our design strategy was empirical, we considered three main types of input into the model. First, measurement of the degree of disease progression at the time of assessment. There is no perfect rating scale for disease progression, our model considered our previously developed scale for sCJD, the MRC Prion Disease Rating Scale, Cognitive and Motor Examination Scales; and the MMSE, clinical category of sCJD and care needs status. Second, prediction of the rate of change of disease progression, which might incorporate inputs from the MRI brain scan, *PRNP* codon 129 genotype, CSF biomarkers and time since disease onset. Third, inputs that relate to the context, age of the patient, sex, and potentially attitudes towards life preserving care at advanced stages, such as preference for the use of tube feeding.

Our primary models predicting survival can be compared to that developed by Llorens *et al*.^18^ which used data from the German Reference Centre for Transmissible Spongiform Encephalopathies to predict 6-month survival from sCJD onset. Both studies used data from national tertiary diagnostic services with similar patient age and sex. Their model of total survival from symptom onset incorporated age, sex, *PRNP* codon 129 genotype, and CSF tau with moderate accuracy (AUC=0.69; 95% CI=0.67–0.71); our models achieved greater performance (AUC 0.91-0.94). We predicted outcomes over a shorter period (10, 30 and 100 days) from date of diagnosis, a more precise time measurement than carer-reported symptom onset which is typically a retrospective estimation, and more useful from the perspective of clinical decision making. Both models included sex and *PRNP* codon 129 genotype. Llorens *et al*. also included age and CSF tau levels, whereas we included a wider range of additional predictors (days since symptom onset, baseline care status, MRC Prion Disease Rating, Motor and Cognitive Scores, MRI abnormalities, MMSE score and clinical phenotype). The improved performance may therefore be in part due to predicting from diagnosis rather than symptom onset, and the wider range of predictors incorporated in the current study.

Our study has some limitations. Given the rarity of sCJD the sample size is large, though from a modelling perspective it is moderate. An external dataset for model validation was not available, and we therefore used leave-one-out cross validation. The sample was particularly limited for patients with early disease and minimal care status, reflecting the difficulty in prompt diagnosis of a rapidly progressive rare disease. Nevertheless, to our knowledge this is the first attempt to prognosticate care status. Due to the limited sample for this secondary subgroup analysis, we did not repeat the variable selection procedure, and instead developed models using the predictors selected for the primary survival models. The pool of potential predictors was determined by what is typically available in the UK at the point of initial diagnosis. Other predictors of potential value, for example serum neurofilament light (NfL), serum total tau, CSF total tau and other biomarkers, and more sophisticated imaging metrics, are only available in the UK as research tests, and were not therefore investigated in this study. A wealth of evidence points to the role of prion strain, or sCJD subtype, in determining clinical phenotype. Unfortunately, there are no direct biomarkers of CJD subtype that can be used in life, although the recent development of an imaging method to infer subtype may prove useful in the future^10^. Further exploration of how to incorporate different imaging abnormalities (eg. atrophy, regions of diffusion abnormality), rather than a simple count, are warranted. As these biofluid and imaging biomarkers move into routine clinical use, they could be evaluated for improving model performance.

Because the apparent annual incidence of sCJD has steadily increased over the last 30 years, it is likely that the disease has not been fully ascertained. The Cohort study did not recruit all patients documented as dying from sCJD in the study period, because either the diagnosis was not made in life, the patient died before we were able to visit, or after 2015, was too advanced in disease stage to be eligible, or the patient or family did not agree to join. It is therefore likely that the study is not fully representative of the theoretical totality of the disease, with diminished ascertainment of the very fastest progressing patients, and those with atypical phenotypes that make the disease hard to diagnose. Additionally, we defined care status in a pragmatic way based on our observations about how care is typically provided to patients with sCJD in the UK. Experiences in other countries may differ, for example regarding the timing and use of formal 24-hour care facilities. Future studies might examine the degree to which models like the one presented here generalize to other countries and populations, with careful consideration about how care status is defined in consideration of local practices.

We should be cautious in assuming that prognostic information about likely survival and care status will necessarily be beneficial to patients and carers, who may have differing views on whether this information is welcome^28^. Indeed, it may even cause harm if this information is shared without careful explanation and compassion. Further pragmatic studies which facilitate clinical implementation of such models are essential. Key outcomes include usability and acceptability to clinicians, patients and carers, and whether the provision of information leads to improved patient outcomes and care provision.

There have been few clinical trials in sCJD relative to other dementias. In part this relates to the rarity of the disease and the challenges evident from rapid progression. Two experimental treatments have currently either entered human use, or are planned for human use: monoclonal antibody therapy and antisense oligonucleotide treatments. The mechanism of actions may require time for the compound to achieve target therapeutic levels in brain tissues, and clinical trials may be less powerful if patients are recruited with a very short prognosis. Prognostic modelling may therefore have value in deciding on eligibility for future clinical trials.

## Conclusions

Among sCJD patients attending a tertiary diagnostic service, routinely collected clinical, imaging and genetic data can be used in combination to accurately predict survival and escalating care status. This has the potential to enhance prognostic decision making during clinical practice and to facilitate the timely provision of support to patients and their families.

## Data Availability

All data produced in the present study are available upon reasonable request to the authors

## Author contributions

JMR contributed to the conception of the work, data analysis, interpretation of data, drafting and revision of the manuscript for intellectual content. DH, KM were involved in the collection and assessment of data and revision of the manuscript for intellectual content. DJL contributed to the conception of the work, interpretation of data, and revision of the manuscript for intellectual content. AN, SM, PR and JC contributed to the conception of the work, obtaining funding, acquisition, interpretation of data, and revision of the manuscript for intellectual content.

## Data Access, Responsibility, and Analysis

JMR and SM had full access to all the data in the study and take responsibility for the integrity of the data and the accuracy of the data analysis. JMR (University of Exeter) conducted and is responsible for the data analysis.

## References

1. Pocchiari M, Puopolo M, Croes EA, et al. Predictors of survival in sporadic Creutzfeldt-Jakob disease and other human transmissible spongiform encephalopathies. Brain. 2004;127(Pt 10):2348–2359.

2. Kortazar-Zubizarreta I, Ruiz-Onandi R, Pereda A, et al. Sporadic Creutzfeldt-Jakob disease with extremely long 14-year survival period. Eur J Neurol. 2021;28(9):2901–2906.

3. Thompson AG, Lowe J, Fox Z, et al. The Medical Research Council prion disease rating scale: a new outcome measure for prion disease therapeutic trials developed and validated using systematic observational studies. Brain. 2013;136(Pt 4):1116–1127.

4. Ford L, Rudge P, Robinson K, Collinge J, Gorham M, Mead S. The most problematic symptoms of prion disease - an analysis of carer experiences. Int Psychogeriatr. 2019;31(8):1181–1190.

5. Appleby BS, Yobs DR. Symptomatic treatment, care, and support of CJD patients. Handb Clin Neurol. 2018;153:399–408.

6. Collinge J. Mammalian prions and their wider relevance in neurodegenerative diseases. Nature. 2016;539(7628):217–226.

7. Brown CA, Schmidt C, Poulter M, et al. In vitro screen of prion disease susceptibility genes using the scrapie cell assay. Hum Mol Genet. 2014;23(19):5102–5108.

8. Collinge J. Variant Creutzfeldt-Jakob disease. Lancet. 1999;354(9175):317–323.

9. Thompson AGB, Anastasiadis P, Druyeh R, et al. Evaluation of plasma tau and neurofilament light chain biomarkers in a 12-year clinical cohort of human prion diseases. Mol Psychiatry. 2021.

10. Bizzi A, Pascuzzo R, Blevins J, et al. Subtype Diagnosis of Sporadic Creutzfeldt-Jakob Disease with Diffusion Magnetic Resonance Imaging. Ann Neurol. 2021;89(3):560–572.

11. Halbgebauer S, Oeckl P, Steinacker P, et al. Beta-synuclein in cerebrospinal fluid as an early diagnostic marker of Alzheimer’s disease. J Neurol Neurosurg Psychiatry. 2020.

12. Llorens F, Kruse N, Schmitz M, et al. Evaluation of alpha-synuclein as a novel cerebrospinal fluid biomarker in different forms of prion diseases. Alzheimers Dement. 2017;13(6):710–719.

13. Franko E, Wehner T, Joly O, et al. Quantitative EEG parameters correlate with the progression of human prion diseases. J Neurol Neurosurg Psychiatry. 2016;87(10):1061–1067.

14. Otto M, Wiltfang J, Schutz E, et al. Diagnosis of Creutzfeldt-Jakob disease by measurement of S100 protein in serum: prospective case-control study. BMJ. 1998;316(7131):577–582.

15. Nihat A, Mok TH, Odd H, et al. Development of novel clinical examination scales for the measurement of disease severity in Creutzfeldt-Jakob disease. MedRxiv. 2020.

16. Staffaroni AM, Kramer AO, Casey M, et al. Association of Blood and Cerebrospinal Fluid Tau Level and Other Biomarkers With Survival Time in Sporadic Creutzfeldt-Jakob Disease. JAMA Neurol. 2019.

17. Sundaram SE, Staffaroni AM, Walker NC, et al. Baseline neuropsychological profiles in prion disease predict survival time. Ann Clin Transl Neurol. 2020;7(9):1535–1545.

18. Llorens F, Rubsamen N, Hermann P, et al. A prognostic model for overall survival in sporadic Creutzfeldt-Jakob disease. Alzheimers Dement. 2020;16(10):1438–1447.

19. Collins GS, Reitsma JB, Altman DG, Moons KG. Transparent Reporting of a multivariable prediction model for Individual Prognosis or Diagnosis (TRIPOD): the TRIPOD statement. Ann Intern Med. 2015;162(1):55–63.

20. Collinge J, Gorham M, Hudson F, et al. Safety and efficacy of quinacrine in human prion disease (PRION-1 study): a patient-preference trial. Lancet Neurol. 2009;8(4):334–344.

21. Hermann P, Appleby B, Brandel JP, et al. Biomarkers and diagnostic guidelines for sporadic Creutzfeldt-Jakob disease. Lancet Neurol. 2021;20(3):235–246.

22. Haaksma ML, Eriksdotter M, Rizzuto D, et al. Survival time tool to guide care planning in people with dementia. Neurology. 2020;94(5):e538–e548.

23. Folstein MF, Folstein SE, McHugh PR. “Mini-mental state”. A practical method for grading the cognitive state of patients for the clinician. J Psychiatr Res. 1975;12(3):189–198.

24. Appleby BS, Appleby KK, Crain BJ, Onyike CU, Wallin MT, Rabins PV. Characteristics of established and proposed sporadic Creutzfeldt-Jakob disease variants. Arch Neurol. 2009;66(2):208–215.

25. Wadsworth JDF, Adamson G, Joiner S, et al. Methods for Molecular Diagnosis of Human Prion Disease. Methods Mol Biol. 2017;1658:311–346.

26. White IR, Royston P, Wood AM. Multiple imputation using chained equations: Issues and guidance for practice. Stat Med. 2011;30(4):377–399.

27. Royston PS W. Multivariable Model-Building: A pragmatic approach to regression analysis based on fractional polynomials for modelling continuous variables. John Wiley & Sons, Ltd; 008.

28. Westeneng HJ, Debray TPA, Visser AE, et al. Prognosis for patients with amyotrophic lateral sclerosis: development and validation of a personalised prediction model. Lancet Neurol. 2018;17(5):423–433.

