## Supplementary material for "Development of Prognostic Models for Survival and Care Status in Sporadic Creutzfeldt-Jakob disease"

**Predicting prognosis and care needs in Sporadic Creutzfeldt-Jakob disease Online Only Supplement**

**Online-only Supplementary Contents**

### eMethods 1. TRIPOD Checklist

| **Section/Topic** | **Item** |  | **Checklist Item** | **Page** |
| --- | --- | --- | --- | --- |
| **Title and abstract** | | | | |
| Title | 1 | D;V | Identify the study as developing and/or validating a multivariable prediction model, the target population, and the outcome to be predicted. | 1 |
| Abstract | 2 | D;V | Provide a summary of objectives, study design, setting, participants, sample size, predictors, outcome, statistical analysis, results, and conclusions. | 3-4 |
| **Introduction** | | | | |
| Background and objectives | 3a | D;V | Explain the medical context (including whether diagnostic or prognostic) and rationale for developing or validating the multivariable prediction model, including references to existing models. | 3-6 |
|  | 3b | D;V | Specify the objectives, including whether the study describes the development or validation of the model or both. | 3-6 |
| **Methods** | | | | |
| Source of data | 4a | D;V | Describe the study design or source of data (e.g., randomized trial, cohort, or registry data), separately for the development and validation data sets, if applicable. | 7 |
|  | 4b | D;V | Specify the key study dates, including start of accrual; end of accrual; and, if applicable, end of follow-up. | 7 |
| Participants | 5a | D;V | Specify key elements of the study setting (e.g., primary care, secondary care, general population) including number and location of centres. | 7 |
|  | 5b | D;V | Describe eligibility criteria for participants. | 7 |
|  | 5c | D;V | Give details of treatments received, if relevant. | 7 |
| Outcome | 6a | D;V | Clearly define the outcome that is predicted by the prediction model, including how and when assessed. | 9 |
|  | 6b | D;V | Report any actions to blind assessment of the outcome to be predicted. | n/a |
| Predictors | 7a | D;V | Clearly define all predictors used in developing the multivariable prediction model, including how and when they were measured. | 8-9 |
|  | 7b | D;V | Report any actions to blind assessment of predictors for the outcome and other predictors. | 7-8 |
| Sample size | 8 | D;V | Explain how the study size was arrived at. | 7 |
| Missing data | 9 | D;V | Describe how missing data were handled (e.g., complete-case analysis, single imputation, multiple imputation) with details of any imputation method. | 10 |
| Statistical analysis methods | 10a | D | Describe how predictors were handled in the analyses. | 10-11 |
|  | 10b | D | Specify type of model, all model-building procedures (including any predictor selection), and method for internal validation. | 10-12 |
|  | 10c | V | For validation, describe how the predictions were calculated. | 11 |
|  | 10d | D;V | Specify all measures used to assess model performance and, if relevant, to compare multiple models. | 11 |
|  | 10e | V | Describe any model updating (e.g., recalibration) arising from the validation, if done. | 11 |
| Risk groups | 11 | D;V | Provide details on how risk groups were created, if done. | n/a |
| Development vs. validation | 12 | V | For validation, identify any differences from the development data in setting, eligibility criteria, outcome, and predictors. | n/a |
| **Results** | | | | |
| Participants | 13a | D;V | Describe the flow of participants through the study, including the number of participants with and without the outcome and, if applicable, a summary of the follow-up time. A diagram may be helpful. | 12 |
|  | 13b | D;V | Describe the characteristics of the participants (basic demographics, clinical features, available predictors), including the number of participants with missing data for predictors and outcome. | 12 |
|  | 13c | V | For validation, show a comparison with the development data of the distribution of important variables (demographics, predictors and outcome). | n/a |
| Model development | 14a | D | Specify the number of participants and outcome events in each analysis. | 12 |
|  | 14b | D | If done, report the unadjusted association between each candidate predictor and outcome. | n/a |
| Model specification | 15a | D | Present the full prediction model to allow predictions for individuals (i.e., all regression coefficients, and model intercept or baseline survival at a given time point). | 12-13 tables |
|  | 15b | D | Explain how to use the prediction model. | 14 |
| Model performance | 16 | D;V | Report performance measures (with CIs) for the prediction model. | 12-13 tables |
| Model-updating | 17 | V | If done, report the results from any model updating (i.e., model specification, model performance). | n/a |
| **Discussion** | | | | |
| Limitations | 18 | D;V | Discuss any limitations of the study (such as nonrepresentative sample, few events per predictor, missing data). | 14 |
| Interpretation | 19a | V | For validation, discuss the results with reference to performance in the development data, and any other validation data. | 14 |
|  | 19b | D;V | Give an overall interpretation of the results, considering objectives, limitations, results from similar studies, and other relevant evidence. | 14-15 |
| Implications | 20 | D;V | Discuss the potential clinical use of the model and implications for future research. | 14-15 |
| **Other information** | | | | |
| Supplementary information | 21 | D;V | Provide information about the availability of supplementary resources, such as study protocol, Web calculator, and data sets. | n/a |
| Funding | 22 | D;V | Give the source of funding and the role of the funders for the present study. | 16 |

### eMethods 2. Expert ratings of potential predictors of sCJD prognosis and care needs

This document summarises the ratings of 25 variables for their likely utility as a predictor of CJD prognosis and care needs. Two clinicians with sCJD assessment expertise (Dr Akin Nihat and Prof Simon Mead) independently rated each variable as 1 (not likely to be useful), 2 (unsure) or 3 (likely to be useful). Potential predictor variables were selected for analysis if they were rated by either clinician as 3, or both clinicians rated the variable as 2. Selected variables are highlighted.

| **Variable Name** | **Details** | **Likely utility rating**  1: Not likely to be useful  2: Unsure  3: Likely to be useful | |
| --- | --- | --- | --- |
|  |  | **AN** | **SM** |
| Sex | 1: Male  2: Female | 1 | 1 |
| Clinical Category Numeric | 1: Visual  2: Ataxic  3: Cognitive  4: Psychiatric/behavioural  5: Sleep/thalamic  6: Stroke-like  7: Classical | 3 | 3 |
| Age at onset |  | 3 | 2 |
| Codon 129 code | 1: MM  2: VV  3: MV | 3 | 3 |
| MRI Atrophy | 1: Present  0: Absent | 2 | 2 |
| MRI Cortical Ribboning | 1: Present  0: Absent | 2 | 1 |
| MRI Pulvinar Sign | 1: Present  0: Absent | 2 | 1 |
| MRI Basal Ganglia Abnormalities | 1: Present  0: Absent | 2 | 1 |
| MRI Thalamic Restriction | 1: Present  0: Absent | 2 | 2 |
| MRI Ordinal | Sum of five MRI categories (0-5) | 3 | 1 |
| EEG Normal? | 1: Abnormal  0: Normal | 1 | 2 |
| EEG PSWCs? | 1: Present  0: Absent | 3 | 2 |
| Non-specific abnormalities | 1: Present  0: Absent | 1 | 1 |
| General Slowing | 1: Present  0: Absent | 1 | 1 |
| EEG Ordinal | 2: PSWCs  1:Slow/non-specific  0: Normal | 2 | 1 |
| CSF White cells | 1: Present at >4 white cells  0: Present at <= 4 white cells or absent | 1 | 1 |
| CSF Protein | 1: Abnormal  0: Normal | 2 | 1 |
| CSF Protein Conc ng/ml |  | 2 | 1 |
| CSF 14-3-3 | 1: Abnormal  0: Normal | 1 | 1 |
| CSF S100b | 1: Abnormal  0: Normal | 2 | 2 |
| CSF S100b Conc ng/ml | 0-13 | 3 | 2 |
| MRC Score /20 | 0-20, intervals of 1 | 3 | 3 |
| Motor Score /100 | 0-100, intervals of 1 | 3 | 2 |
| Cognitive Score /100 | 0-100, intervals of 1 | 3 | 2 |
| MMSE /30 | 0-30, intervals of 1 | 3 | 1 |

### eTable 1. Baseline characteristics of study participants (N = 537)

|  | **10-day mortality** | | **30-day mortality** | | **100-day mortality** | |
| --- | --- | --- | --- | --- | --- | --- |
|  | Survived  N = 410 | Died  N = 127 | Survived  N = 244 | Died  N = 293 | Survived  N = 123 | Died  N = 414 |
| Age in years, median (IQR) | 66.82 (10.04) | 68.68 (11.68) | 65.05 (11.16) | 68.72 (10.85) | 64.76 (11.46) | 67.93 (10.52) |
| Female, N (%) | 218 (53.17) | 62 (48.82) | 122 (50.00) | 158 (53.92) | 68 (55.28) | 212 (51.21) |
| Days since onset, median (IQR)* | 147.50 (179.00) | 77.00 (61.00) | 190 (187.5) | 85 (86.00) | 232.00 (162.00) | 95.00 (119.00) |
| Nursing home / hospice care, N (%) | 93 (22.68) | 105 (82.68) | 25 (10.25) | 173 (59.04) | 12 (9.76) | 186 (44.93) |
| Codon 129 polymorphism* |  |  |  |  |  |  |
| MM, N (%) | 149 (36.34) | 99 (77.95) | 57 (23.36) | 191 (65.19) | 57 (23.36) | 191 (65.19) |
| VV, N (%) | 119 (29.02) | 17 (13.39) | 68 (27.87) | 68 (23.21) | 68 (27.87) | 68 (23.21) |
| MV, N (%) | 142 (34.63) | 11 (8.66) | 119 (48.77) | 34 (11.60) | 119 (48.77) | 34 (11.60) |
| MRC score, median (IQR)* | 9.00 (9.00) | 1.00 (3.00) | 11.00 (9.00) | 2.00 (7.00) | 13.00 (9.00) | 4.00 (9.00) |
| Motor score, median (IQR)* | 38.76 (38.02) | 0.00 (6.00) | 47.97 (25.84) | 7.00 (33.28) | 53.40 (22.49) | 18.70 (39.43) |
| Cognitive score, median (IQR)* | 28.26 (58.80) | 0.00 (0.00) | 45.94 (45.85) | 0.00 (11.00) | 48.14 (44.85) | 1.00 (38.00) |
| EEG PSWCs, N (%)* | 88 (21.46) | 65 (51.18) | 24 (9.84) | 129 (44.03) | 10 (8.13) | 143 (34.54) |
| CSF s100b abnormality, N (%)* | 226 (55.12) | 65 (51.18) | 125 (51.23) | 166 (56.66) | 64 (52.03) | 227 (54.83) |
| MRI abnormalities, median (IQR)* | 2 (2.00) | 2 (2.00) | 2.00 (2.00) | 2.00 (2.00) | 2.00 (2.00) | 2.00 (2.00) |
| MMSE score, median (IQR)* | 2 (15.00) | 0 (0.00) | 10.00 (19.00) | 0.00 (.00) | 11.00 (20.00) | 0.00 (8.00) |
| Clinical phenotype* |  |  |  |  |  |  |
| Classical, N (%) | 138 (33.66) | 49 (38.58) | 67 (27.46) | 120 (40.96) | 32 (26.02) | 155 (37.44) |
| Cognitive, psychiatric, behavioural, N (%) | 125 (30.49) | 42 (33.07) | 90 (36.89) | 77 (26.28) | 55 (44.72) | 112 (27.05) |
| Other, N (%) | 147 (35.85) | 36 (28.35) | 87 (35.66) | 96 (32.76) | 36 (29.27) | 147 (35.51) |

Notes: IQR = Interquartile Range, MRC = Medical Research Council, EEG = Electroencephalography, PSWC = Periodic Sharp Wave Complexes, CSF = Cerebrospinal Fluid, MRI = Magnetic Resonance Imaging, MMSE = Mini-Mental State Examination. Clinical phenotype ‘other’ includes visual, ataxic, sleep/thalamic and stroke-like phenotypes. *There was missing data on this variable. Descriptive statistics are reported for imputed data.

### eTable 2. Final prediction models for 10-, 30- and 100-day mortality

|  | **10-day mortality** | | | **30-day mortality** | | | **100-day mortality** | | |
| --- | --- | --- | --- | --- | --- | --- | --- | --- | --- |
|  | OR | *p* | 95% CI | OR | *p* | 95% CI | OR | *p* | 95% CI |
| Male | 2.48 | 0.006 | 1.29 - 4.77 | 1.15 | 0.615 | 0.67 - 1.96 | 1.92 | 0.030 | 1.07 - 3.45 |
| Days since onset | 1.00*^a^* (p1) | 0.015 | 1.00 - 1.00 | 0.41 *^c^* (p1) | <0.0001 | 0.29 - 0.58 | 1.14*^e^* (p1) | 0.000 | 1.08 - 1.20 |
|  | 1.16 *^a^* (p2) | 0.003 | 1.05 - 1.27 |  |  |  |  |  |  |
| Nursing home / hospice care | 4.02 | <0.0001 | 1.87 - 8.66 | 3.25 | 0.001 | 1.66 - 6.40 | 1.15 | 0.759 | 0.47 - 2.83 |
| Codon 129 polymorphism |  |  |  |  |  |  |  |  |  |
| MM (versus MV) | 2.57 | 0.124 | 0.77 - 8.60 | 6.65 | <0.0001 | 3.02 - 14.68 | 4.33 | 0.000 | 1.97 - 9.48 |
| VV (versus MV) | 1.65 | 0.437 | 0.46 - 5.89 | 2.61 | 0.014 | 1.21 - 5.60 | 5.32 | 0.000 | 2.51 - 11.25 |
| Motor score | 0.97 | 0.03 | 0.94 - 1.00 | 0.00 *^d^* (p1) | 0.002 | 0.00 - 0.04 | 0.97 | 0.016 | 0.95 - 0.99 |
| Cognitive score | 6.9*^b^* (p1) | 0.002 | 2.06 - 23.27 | 1.00 | 0.938 | 0.98 - 1.02 | 1.00 | 0.654 | 0.98 - 1.03 |
|  | 1.17*^b^* (p2) | 0.002 | 1.06 - 1.29 |  |  |  |  |  |  |
| MRC score | 1.07 | 0.398 | 0.91 - 1.26 | 0.96 | 0.485 | 0.87 - 1.07 | 0.88 | 0.024 | 0.79 - 0.98 |
| MRI abnormalities | 1.29 | 0.093 | 0.96 - 1.74 | 1.12 | 0.347 | 0.88 - 1.42 | 1.11 | 0.400 | 0.87 - 1.43 |
| MMSE score | 1.15 | 0.028 | 1.02 - 1.31 | 0.97 | 0.202 | 0.92 - 1.02 | 1.01 | 0.741 | 0.96 - 1.06 |
| Clinical phenotype |  |  |  |  |  |  |  |  |  |
| Classical (versus other) | 1.75 | 0.326 | 0.57 - 5.39 | 1.81 | 0.111 | 0.87 - 3.76 | 1.01 | 0.974 | 0.46 - 2.22 |
| Cognitive, psychiatric, behavioural (versus other) | 1.15 | 0.816 | 0.35 - 3.82 | 0.61 | 0.251 | 0.27 - 1.41 | 0.44 | 0.047 | 0.20 - 0.99 |

Notes: OR = Odds Ratio, CI = Confidence Interval; MRC = Medical Research Council; MRI =Magnetic Resonance Imaging; MMSE = Mini-Mental State Examination; Clinical phenotype ‘other’ includes visual, ataxic, sleep/thalamic and stroke-like phenotypes; The following second degree fractional polynomial terms were used: *^a^*(-2, -1); *^b^* (-0.5, -0.5). The following first degree fractional polynomial terms were used: *^c^*(0), *^d^*(3). *^e^*(-1).

### eTable 3. Leave-one-out cross validation of models predicting 10- 30- and 100-day mortality in sCJD patients (N=537)

|  | **10-day**  **mortality** | **30-day**  **mortality** | **100-day**  **mortality** |
| --- | --- | --- | --- |
| Area Under the Curve | 0.91 | 0.90 | 0.89 |
| Correctly classified (%)* | 85.9% | 81.8% | 84.5% |
| Sensitivity* | 72.4% | 84.0% | 91.8% |
| Specificity* | 90.0% | 79.1% | 60.2% |
| Positive Predictive Value* | 69.2% | 82.8% | 88.6% |
| Negative Predictive Value* | 91.3% | 80.4% | 68.5% |

### eTable 4. Final prediction models for 10-, 30- and 100-day increased care needs

|  | **10-day increased care needs** | | | **30-day increased care needs** | | | **100-day increased care needs** | | |
| --- | --- | --- | --- | --- | --- | --- | --- | --- | --- |
|  | OR | *p* | 95% CI | OR | *p* | 95% CI | OR | *p* | 95% CI |
| Male | 2.40 | 0.257 | 0.53 - 10.88 | 2.24 | 0.183 | 0.68 - 7.33 | 156.91 | 0.001 | 7.50 - 3282.85 |
| Days since onset | 1.00*^a^* (p1) | 0.035 | 1.00 - 1.00 | 1.00 *^a^* (p1) | 0.022 | 1.00 - 1.01 | 1.01 *^a^* (p1) | 0.104 | 1.00 - 1.02 |
| Nursing home / hospice care | 0.41 | 0.361 | 0.06 - 2.80 | 0.19 | 0.019 | 0.05 - 0.77 | 0.22 | 0.132 | 0.03 - 1.58 |
| Codon 129 polymorphism |  |  |  |  |  |  |  |  |  |
| MM (versus MV) | 2.57 | 0.379 | 0.31 - 20.96 | 1.32 | 0.734 | 0.27 - 6.41 | 0.43 | 0.499 | 0.04 - 4.91 |
| VV (versus MV) | 5.99 | 0.045 | 1.04 - 34.45 | 2.91 | 0.118 | 0.76 - 11.12 | 1.28 | 0.801 | 0.19 - 8.72 |
| Motor score | 1.00 | 0.892 | 0.95 - 1.06 | 0.99 | 0.752 | 0.95 - 1.04 | 0.96 | 0.232 | 0.89 - 1.03 |
| Cognitive score | 1.00 | 0.908 | 0.96 - 1.04 | 1.00 | 0.896 | 0.96 - 1.04 | 0.96 | 0.266 | 0.89 - 1.03 |
| MRC score | 0.86 | 0.280 | 0.65 - 1.13 | 0.84 | 0.125 | 0.67 - 1.05 | 0.60 | 0.045 | 0.37 - 0.99 |
| MRI abnormalities | 1.23 | 0.509 | 0.66 - 2.28 | 1.28 | 0.329 | 0.78 - 2.11 | 1.45 | 0.388 | 0.62 - 3.38 |
| MMSE score | 0.94 | 0.165 | 0.85 - 1.03 | 1.01 | 0.839 | 0.93 - 1.09 | 1.04 | 0.59 | 0.90 - 1.19 |
| Clinical phenotype |  |  |  |  |  |  |  |  |  |
| Classical (versus other) | 0.48 | 0.392 | 0.09 - 2.60 | 0.14 | 0.009 | 0.03 - 0.60 | 0.06 | 0.032 | 0.00 - 0.78 |
| Cognitive, psychiatric, behavioural (versus other) | 0.26 | 0.109 | 0.05 - 1.34 | 0.13 | 0.009 | 0.03 - 0.60 | 0.01 | 0.007 | 0.00 - 0.25 |

Notes: OR = Odds Ratio, CI = Confidence Interval; MRC = Medical Research Council; MRI =Magnetic Resonance Imaging; MMSE = Mini-Mental State Examination; Clinical phenotype ‘other’ includes visual, ataxic, sleep/thalamic and stroke-like phenotypes; *^a^*A first degree fractional polynomial term was used (-2).

### eTable 5. Leave-one-out cross validation of models predicting 10- 30- and 100-day increased care needs in sCJD patients (N=113)

|  | **10-day**  **increased care needs** | **30-day**  **increased care needs** | **100-day**  **increased care needs** |
| --- | --- | --- | --- |
| Area Under the Curve | 0.72 | 0.77 | 0.86 |
| Correctly classified (%)* | 81.4% | 70.8% | 85.8% |
| Sensitivity* | 30.00 | 67.27 | 90.91 |
| Specificity* | 92.47 | 74.14 | 68.00 |
| Positive Predictive Value* | 46.15 | 71.15 | 90.91 |
| Negative Predictive Value* | 86.00 | 70.49 | 68.00 |
